# Performance and Thermal Perceptions of Runners Competing in the London Marathon: Impact of environmental conditions

**DOI:** 10.1101/2020.05.22.20110015

**Authors:** Tim Vernon, Alan Ruddock, Maxine Gregory

## Abstract

**Objectives:** The 2018 Virgin Money London Marathon (2018VMLM) was the hottest in the race’s 37 year history, the major aims of this research were to 1) survey novice-mass participation marathoners to examine the thermal demands of the extreme weather event; and 3) investigate the effect of the air temperature on finish times.

**Methods:** A mixed methods design involving the collection of survey data (n = 364; male = 63, female = 294) and secondary analysis of environmental and marathon performance (676,456 finishers) between 2009 and 2018 was used.

**Results:** The 2018VMLM was hotter than the mean of all other marathons (*P <* 0.05); mean finishing time was slower than the mean of all other London Marathons (*P <* 0.05); there were positive correlations between maximum race-day temperature and finish time for mass-start participants and championship runners, and the difference in maximum race day temperature and mean maximum daily temperature for the 60 days prior to the London Marathon (*P <* 0.05). 23% of surveyed participants classified their thermal sensation and comfort as ‘warm’, ‘hot’ or ‘very hot’, with 68% ‘comfortable’ during training compared with a peak of 95% feeling ‘warm’, ‘hot’ or ‘very hot’ and 77% ‘uncomfortable’ or ‘very uncomfortable’ during 2018VMLM.

**Conclusion:** Organisers should use temperature forecasting and plan countermeasures such as adjusting the start time of the event to avoid high-temperatures; help runners predict finish time and adjust pacing strategies accordingly and provide safety recommendations for participants at high risk time-points as well as cooling strategies.

## INTRODUCTION

Over one million runners participate in marathons annually[1], with the Virgin Money London Marathon, attracting around 40,000 participants. The demands of marathon running are considerable irrespective of performance standard, the energy expenditure of female and male runners finishing between 2 and 4 hours is within the region of 2000 to 2800 kcal, placing extensive strain on metabolic, cardiorespiratory, thermophysiological, mechanical and perceptual regulatory systems[2, 3]. Such demands are intensified in hot and humid conditions where finishing times are impaired [4,5,6], and in-race withdrawals increase when air temperature exceeds 20°C [6].

Substantial evidence supports the increased likelihood of extreme global weather events in the forthcoming decades, including periods of unseasonably cold and hot weather. In February and March 2018, the UK experienced two severe winter weather events with unseasonably low temperatures and significant snowfall [7], For runners preparing for the Virgin Money London Marathon these low temperatures posed logistical training demands (icy roads) and altered perceptual and physiological responses, potentially impairing preparation for a spring marathon. These weather events were followed by unseasonably high temperatures between 18^th^ and 22^nd^ of April, including the UK’s warmest April day since 1949 at 29.1°C, coinciding with the hottest London Marathon on record of 24.1°C, with many mass participation runners still on course when this temperature was recorded. This data, however, does not capture the potential higher temperatures within the micro-climate on course, whereby runners grouped nearby can experience increases in mean radiant temperature of 2°C, coupled with decreases in radiative and convective heat transfer increasing autonomic thermoregulatory challenge. [8] It is also unclear how thermal sensation and comfort, which are integral to behavioural thermoregulation and dominant factors that dictate perceived exertion in hot environments [9] are influenced in these specific environments.

In the final weeks and days of marathon preparation runners were faced with 2 polarised, and conflicting extreme weather events, given, that these extreme weather events are likely to increase in either frequency and/or severity, and the thermophysiological and perceptual demands of marathon running are extensive and increase the risk to the short and long-term health and wellbeing of participants, the aims of this research were to 1) identify historical temperature data for all London Marathons to place the extreme weather events of 2018 in context; 2) survey a sample of novice-mass participation marathoners who started the 2018 Virgin Money London Marathon to examine the thermal demands of the extreme weather event on race day and 3) investigate the effect of the extreme weather events on finish time in both mass participation and championship runners. This investigation is a first and warranted on the basis of the aforementioned risks to health and wellbeing, the addition to our knowledge and understanding of perceptual demands of mass participation runners in the Virgin Money London Marathon, and identification of potential areas whereby participants and race organisers might seek to change their practice in preparation for, or on the day of the event when high-temperatures are forecast.

## METHOD

### Research design

A mixed-methods design involving the collection of survey data and the secondary analysis of environmental and marathon performance data was used to examine the experiences of and the effects of air temperature on participants running a marathon in the heat. The local ethics committee approved the study (ER6896994), all participants provided digital informed consent, and the investigation was conducted in accordance with the Declaration of Helsinki (7th revision).

### Participants

The finish times of 676,456 finishers (male, 450,071; female, 226,385) of the London Marathon from 2001 to 2019, and 180 male London Marathon Championship runners from the years 2009 through 2019 were extracted from the official website of the Virgin Money London Marathon[10] and the marathon archives website[11].

Participants (n = 364; male = 63, female = 294; Age = 41.4 ± 8.3 years; Mass = 72.2 ± 19.9 kg; Stature = 168.6 ± 9.5 cm) from the 2018 Virgin Money London Marathon completed an online survey relating to their expectations and experiences of the event, including expected and actual finish time, perception of temperature, and thermal comfort during the Marathon and during training over the winter months. The survey was distributed via social media platforms and hosted on the Key Survey platform (www.keysurvey.co.uk). The group were mostly inexperienced with the 2018 Virgin Money London Marathon being the first marathon for 63% of runners, 33% of runners had participated in between two and five marathons and only 4% having run more than six marathons. Retrospective analysis of championship runners performance times from 2010 to 2019 (Championship runners defined as athletes invited to participate by the organising committee) were also investigated in relation to indices of temperature.

### Data analysis

Hourly temperature data (°C) were acquired from the UK’s National Meteorological Service (the Met Office) for the date of the London Marathon and the preceding 60 days for the years 1981 to 2019. These data were recorded as per the Met Office standards at a meteorological station located in St James’s Park London, chosen for its central location on the London Marathon route.

Data were processed, plotted and analysed alongside marathon performance data using customised Microsoft Excel Software (Microsoft Corp, 2013) and SPSS (Version 24.0. Armonk, NY: IBM Corp) and assumptions for parametric statistical analyses were performed. Pearson Product Moment Correlation was used to investigate the relationship between maximum race day air temperature and the differential temperature between race day temperature and average maximum temperature for the previous 60 days on the average London Marathon finish time between 2001 and 2019 for the mass participation runners and between 2009 and 2019 for the Championship athletes. Thresholds of 0.1, 0.3, and 0.5 for small, moderate, and large correlations[12] and 0.7 and 0.9 for very large and extremely large correlations[13] were used to interpret relationships between variables. Independent t-tests were performed using SPSS to assess the difference in London Marathon finish times, air temperature and expected versus actual finish time from survey respondents. Statistical significance was set at *P <* 0.05. Thermal perception and comfort data were analysed using SPSS (Version 24.0. Armonk, NY: IBM Corp) using a Friedman test and Post Hoc Man Whitney Test, statistical significance was set at *P* < 0.05.

## RESULTS

### Environmental data

The maximum air temperature recorded by the Met Office on the day of the 2018 Virgin Money London Marathon was 24.1°C, hotter than the mean maximum race day temperature for every other year (15.2 ± 3.3°C). The maximum daily temperature in the 60 days before the 2018 race day was also lower than previous years (11.3 ± 6.3°C vs. 12.1 ± 1.6°C; *P <* 0.01), demonstrated in figure 1 which also shows three extreme weather events of 2018 (a, b and c).

**Figure 1:**
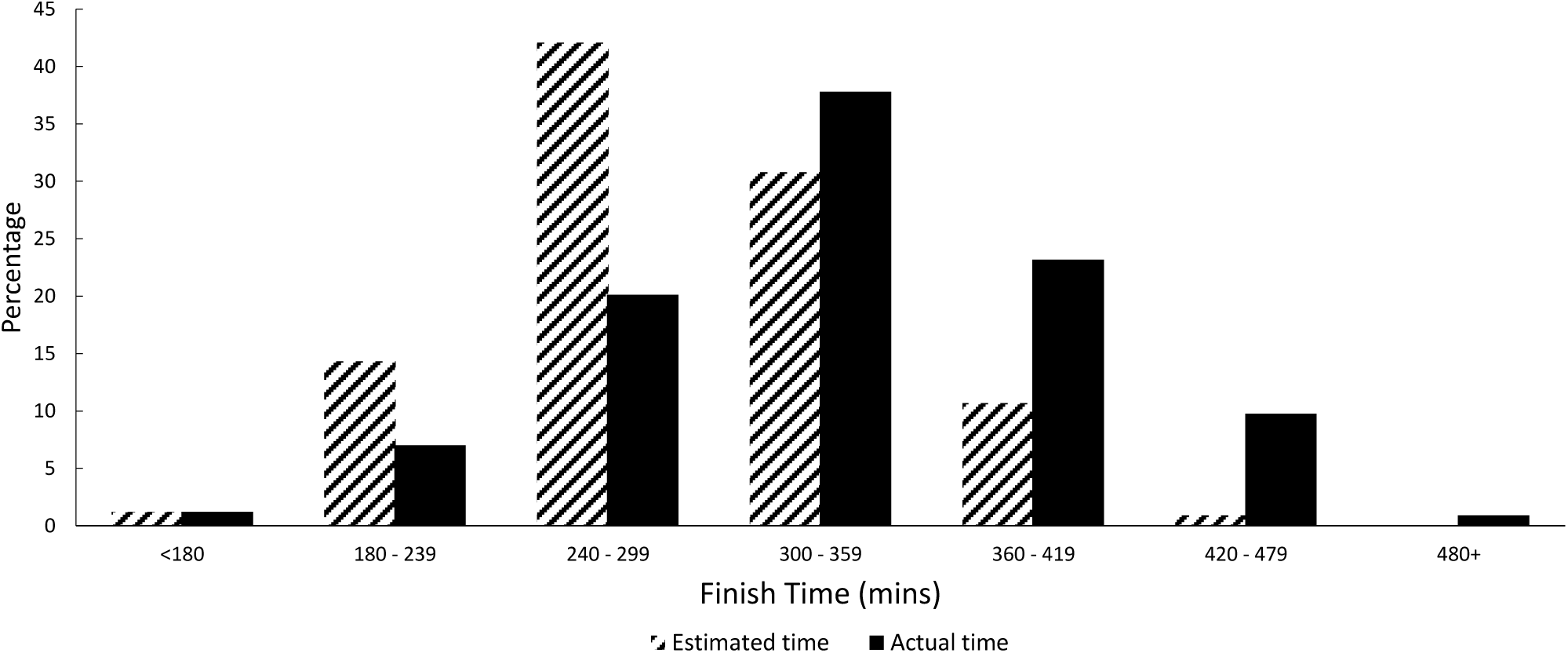
Maximum daily air temperature (°C) in the 60 days leading up to London Marathon race day for 2018 and mean ± SD across all other years 1981 – 2017 and 2019.

### Finish time

The mean finish time of survey respondents for the 2018 VMLM (Figure 2) was 337 ± 51 minutes, slower than the mean finish time for the 2018 Virgin Money London Marathon of 290 ± 64 minutes and 47 ± 30 minutes or 14% slower than participants estimated time (*P <* 0.05).

**Figure 2:**
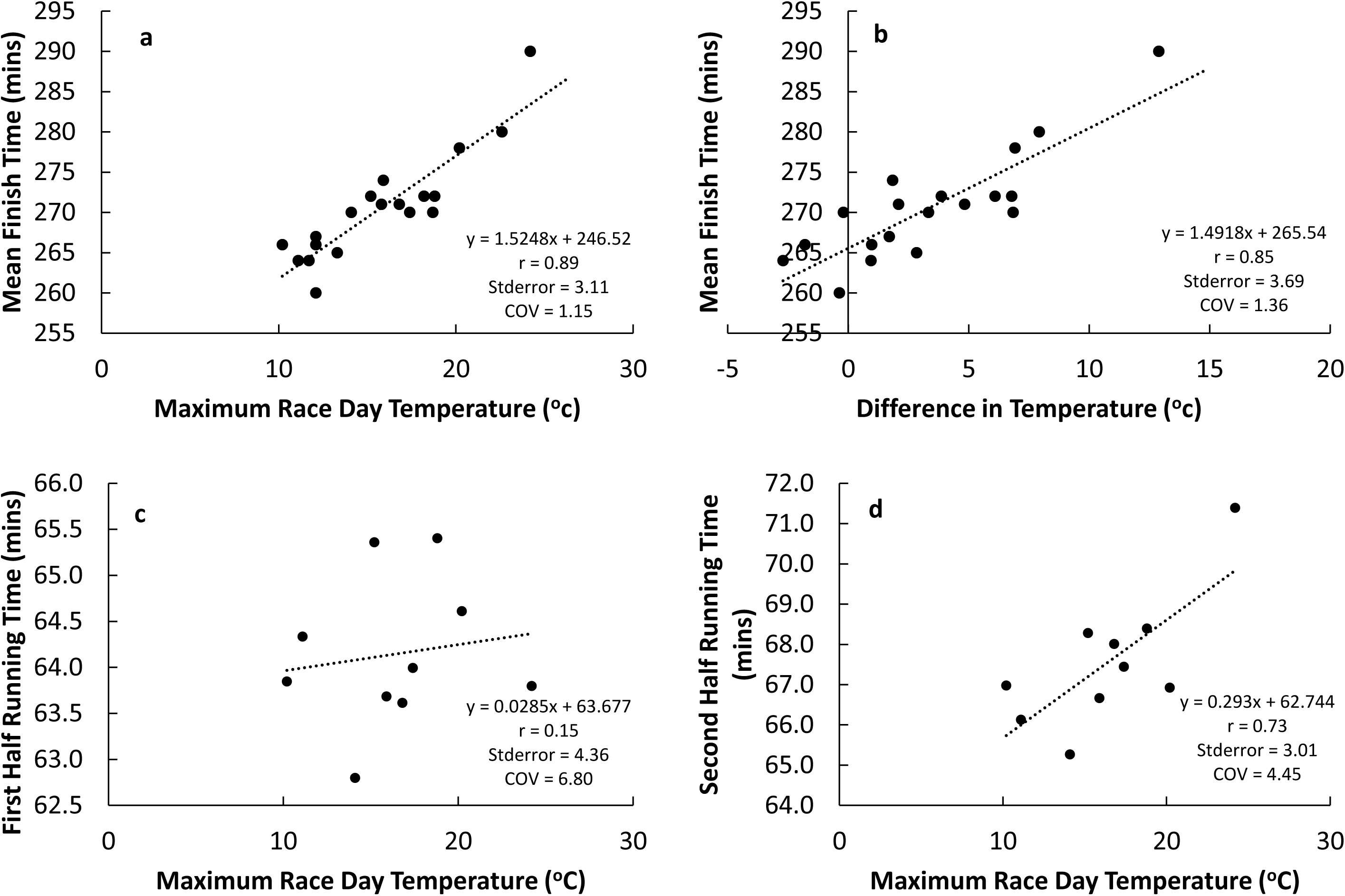
Estimated versus actual finish times in the 2018 VMLM for respondents of the study survey.

For those who had run marathon’s previously, the mean finish time for the 2018 VMLM was 40 ± 27 minutes slower than their previous best marathon time (317 ± 71 mins vs 278 ± 60 mins; *P <* 0.05). Out of 101 runners who had previously run a marathon, one runner ran faster in 2018 VMLM by approximately 1 minute.

The total field (*n* = 40,255) mean completion time for the VMLM 2018 was 20 minutes slower than the mean of all other years between 2001 and 2019 (290 minutes vs 270minutes). There was a very large positive correlation (*r* = 0.89, *P <* 0.05) between maximum race day temperature and mean completion time for mass participation runners (*n* = 676,456) (Figure 3a); and very large positive correlation (*r =* 0.85, *P <* 0.05) between the mean maximum daily temperature for the 60 days before the London Marathon and mean completion time for mass participation runners (Figure 3b).

For Championship runners, the effect of temperature during the first half of the marathon was negligible (*r* = 0.15, *P >* 0.05) (Figure 3c). During the second half of the race, there was a strong positive correlation (*r =* 0.73, *P <* 0.05) between the maximum race day temperature and running time (Figure 3d). This culminates in a strong positive correlation (*r* = 0.65, *P <* 0.05) between the maximum race day temperature and mean marathon finish time for Championship Athletes.

**Figure 3:**
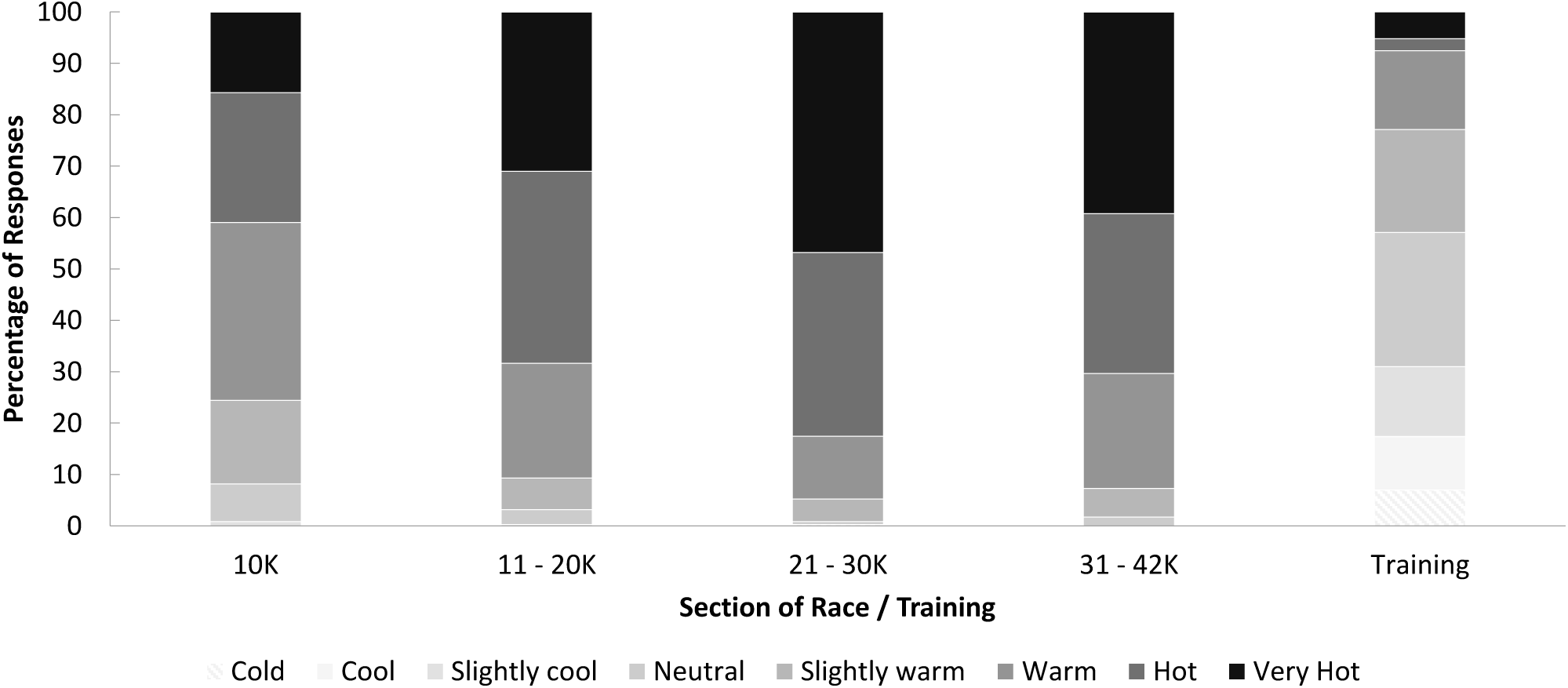
a) Relationship between maximum race day temperature and mean finish time in the London Marathon 2001 to 2019; b) Relationship between the difference in maximum race day temperature and mean-maximum daily temperature during the 60 days prior to race day on mass participation and mean finish time in the London Marathon 2001 to 2019; c) Relationship between maximum race day temperature and championship runners mean first half times in the London Marathon 2010 to 2019; d) Relationship between maximum race day temperature and championship runners mean second half times in the London Marathon 2010 to 2019.

### Perception of heat

To assess thermal sensation, participants were asked “how hot did you feel during the marathon, and how does this compare with your training?” they reported feeling hotter during the VMLM 2018 compared with a typical training run (Figure 4). Runner’s thermal sensation increased as the race progressed with 76% feeling “warm” to “very hot” in the first 10 km, increasing to 91% of participants after the first 10 km. The percentage of participants who felt “warm” to “very hot” during a typical training run was 23%.

**Figure 4:**
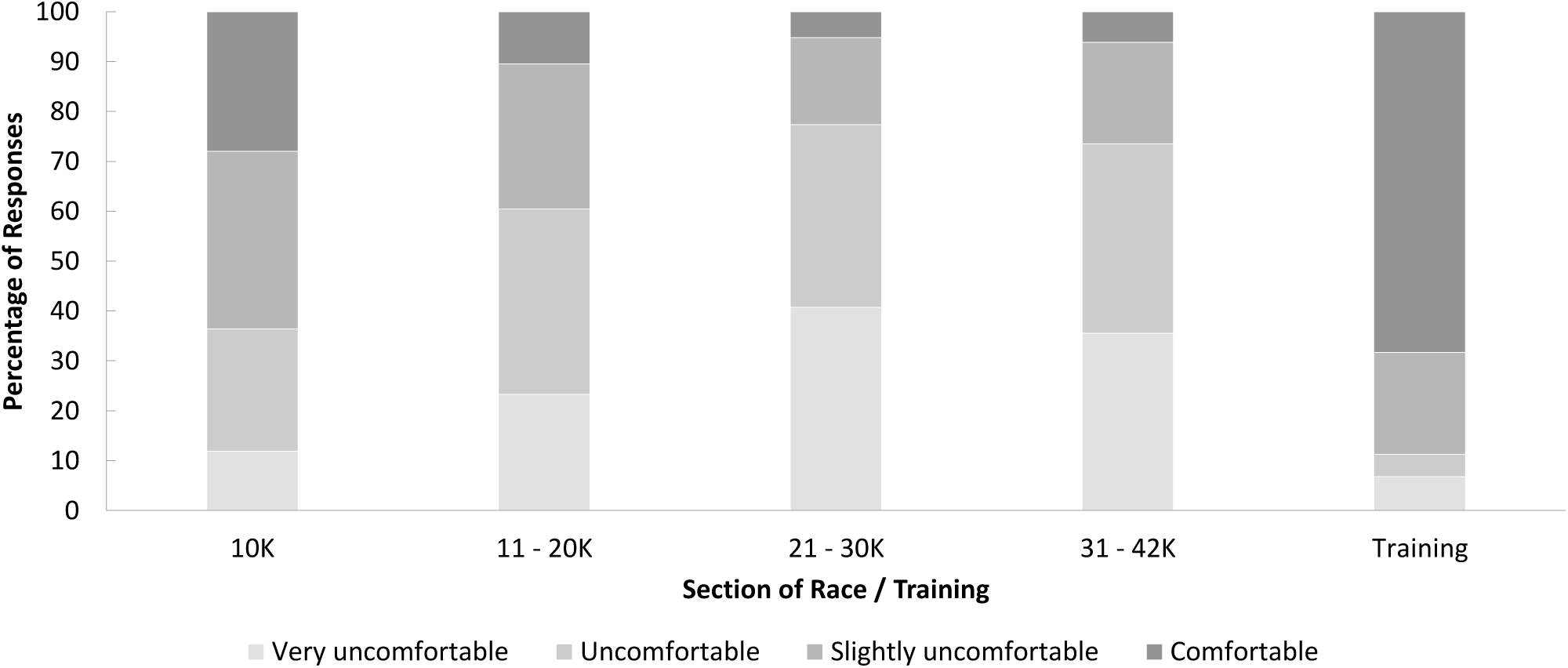
Thermal sens ation during the VMLM 2018 and a typical training session

When asked how comfortable runners were with the temperature during the different stages of the VMLM 2018 and a typical training run (Figure 5), the percentage of runners feeling “uncomfortable” or “very uncomfortable” increased from 37% during the first 10 km to 77% between kilometres 21 and 30. The majority of participants felt comfortable during a typical training run, 11% of respondents felt “uncomfortable” or “very uncomfortable” with the temperature during a typical training run.

**Figure 5:**
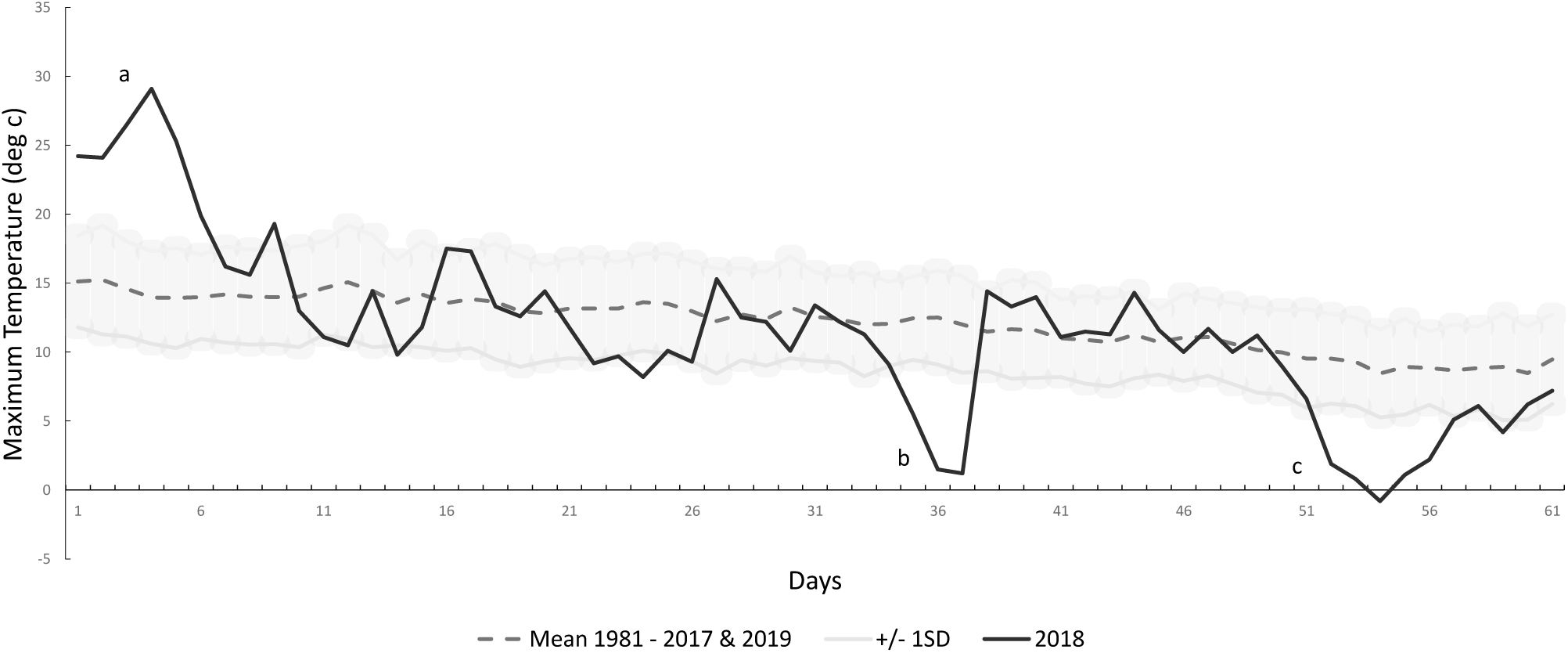
Thermal comfort during VMLM 2018 and a typical training session

## DISCUSSION

The aims of this research were to 1) identify historical temperature data for all London Marathons to place the extreme weather events of 2018 in context; 2) survey a sample of participants who started the 2018 Virgin Money London Marathon to examine the thermoperceptual demands of the extreme weather event on race day and 3) investigate the effect of the extreme weather events on finish time in both mass participation and championship runners. To our knowledge, this investigation is the first to analyse historical weather data of the Virgin London Marathon in context to previous weather data, assess the thermoperceptual demands and determine the effect of ambient temperature on the finish time of runners. The findings from this investigation are that; 1) the 2018 Virgin Money London Marathon was hotter than the mean of all other London Marathons; 2) the 2018 Virgin London Marathon mean finishing time was slower than the mean of all other London Marathons; 3) in accordance with the aforementioned major findings we found a positive correlation between maximum race day temperature and finish time for mass participants, whereby a hotter temperature was related to a slower finish time; 4) we also found a positive correlation between the difference in maximum race day temperature and the mean maximum daily temperature for the 60 days prior to the London Marathon, whereby a hotter race day temperature compared to mean training temperature resulted in a slower marathon finish time; 5) championship runners demonstrated similar relationships between maximum daily temperature and finish time, whereby hotter temperatures were associated with a slower finishing time; 6) only 23% of surveyed participants classified their thermal sensation as ‘warm’, ‘hot’ or ‘very hot’ during their training period compared with a peak of 95% of participants feeling ‘warm’, ‘hot’ or ‘very hot’ during the 2018 Virgin Money London Marathon; and 7) 68% of surveyed participants felt ‘comfortable’ with their body temperature during training whereas a peak of 77% felt “uncomfortable or ‘very uncomfortable’ during the 2018 Virgin Money London Marathon.

Our findings that the 2018 Virgin London Marathon was hotter and finish time was slower than previous London Marathon’s are in accordance with previous research indicating that finish times are slower in hot conditions compared with cooler conditions[4-6]. Predictions of performance impairment based on ambient temperature suggest a decrease in finish time between 1.5 to 2% for every 5°C increase in ambient temperature above 10 to 12°C,[14,15] our data which relates specifically to our sample from the London Marathon, and our sample with mean finish times between 260 and 290 minutes, suggests there is approximately a 2.8% decrease in finish time for every 5°C increase in ambient temperature. A similar relationship (approximately 2.6%) is evident when considering the difference between maximum race day temperature and the mean temperature 60 days before the London Marathon. This knowledge is important because it could be used before any London marathon to adjust pacing strategies by taking into account the St. James Park weather forecast. For example, a runner with an expected finish time of 3 hours 59 minutes derived from training data in the 60 days before the London marathon at a mean temperature of 12°C, would need to adjust their performance time by approximately 5.6%, from a mean pace of 5:40 min/km to 5:59 min/km if the expected maximum daily temperature at St. James Park was forecast to be 22°C.

The extreme warm weather event recorded and reported by the Met Office in the days before the VMLM 2018 meant that the 24.1°C air temperature experienced by the participants was the hottest in the race’s 37-year history. This was hotter than the mean maximum race day temperature for all other London Marathon events. The two cold extreme weather events reported by the Met Office also impacted the daily temperatures in the lead up to the 2018 VMLM, with mean maximum temperature over this crucial training period being lower than in the lead up to any other London Marathon. The net effect was a larger than average differential between the temperature on race day compared to that during the training period in 2018 than in any other London Marathon year. Based on the 2018 VMLM temperature data, the expected performance decrement according to the differential temperature regression equation (figure 3b) predicts a performance impairment of 6.6%, however, our sample of runners experienced a 14% decrement in finish time compared to their estimated finish time derived from training data. This discrepancy might be explained by the pacing strategies employed by respondents who attempted to start the race at their expected race pace, rather than adjust their pace to offset the impact of higher than expected temperatures on thermophysiological, energetic and perceptual demands. This discrepancy may be further accounted for by increases in microclimate temperature, in particular an increased relative humidity caused by evaporative heat loss through sweating and respiration, in our sample of runners who were likely running together in tight groups with limited airflow.

Only 20% of surveyed participants classified their thermal sensation as ‘warm’, ‘hot’ or ‘very hot’ during their training period compared with a peak of 80% of participants feeling ‘hot’ or ‘very hot’ during the 2018 Virgin Money London Marathon. Moreover, 70% of surveyed participants felt their thermal comfort during training was ‘comfortable’ whereas a peak of around 80% felt “uncomfortable and ‘very uncomfortable’ during the VMLM 2018. During exercise in hot environments the two key inputs directly related to RPE are the rate of increase and/or magnitudes of thermal sensation, thermal comfort and cardiovascular [9], Initial predictions regarding intensity of exercise are primarily made based upon skin temperature, which has a large influence on ratings of thermal comfort and thermal sensation followed by cardiovascular strain and ventilatory rate (breathlessness). It is possible that at the start of the marathon, when ratings of thermal sensation and comfort were similar to training, RPE, thus pacing, was also the same. Ratings of thermal sensation and comfort are important from both thermoregulatory and homeostatic perspectives as increases in both likely reflect an increase in whole-body thermophysiological demand, in particular body temperature and significant cardiovascular challenges to maintain homeostasis. Approximately 70% of respondents felt ‘hot’ from 11 – 20 km, peaking at 80% between 21 – 30 km, compared to 40% in the first 10 km, we do not have medical records accompanying this data, however, previous research conducted on the 2007 London Marathon, which was also relatively hot (air temperature = 19.1°C) compared to previous years (air temperature = 11.6°C) reported a mean finish time 17 min slower than previous years. In the 2007 London Marathon 5032 runners were treated by St. John’s Ambulance, there were 73 hospitalisations, 6 cases of severe electrolyte imbalance and 1 death (hyponatreamia) compared to the 2008 London Marathon (air temperature = 9.9°C) where the number of runners treated was 4000. It is not possible to isolate heat-related issues within these figures[16]; however, there is an association between the percentage of withdrawal’s from races and increasing air temperature over 15°C[17], Given the several risk factors associated with exercise in hot environments[18] global recommendations for event cancellation based on Wet Bulb Globe Temperature should be considered as guide [19]. The American College of Sports Medicine[20] suggest the acceptable upper limit for competition as a WBGT of 30.1°C, whereas Roberts[16] suggests that marathons for non-elite runners should be cancelled if the WBGT exceeds 20.5°C at the start of the race, this assumes that races starting in the morning increase in temperature throughout the race. These recommendations, however, do not take into account acclimation state of runners and given the unseasonably low temperatures preceding the 2018 VMLM it is reasonable to assume that the majority of mass participation runners would not have had the opportunity to prepare in environmental conditions similar to race day. Indeed, our analysis suggests a strong relationship between the difference in maximum race day temperature and the mean maximum daily temperature for the 60 days before the London Marathon (Figure 3b).

### Practical Recommendations

Organisers of marathons, particularly in the United Kingdom and Northern Europe should follow existing guidance on exercise in hot environments[19] and specifically; 1) use temperature forecasting and plan countermeasures such as adjusting the start time of the event to avoid high-temperatures; 2) identify ‘at-risk’ participants, such as those who are unacclimated; 3) help runners predict finish time and adjust pacing strategies accordingly using our data analysis; 4) use data regarding thermal perception to provide recommendations for participants as to high-risk time-points of events to help alleviate discomfort; 5) use thermal perception data to provide participants with on-course cooling and fluid guidelines; 6) implement cooling strategies such as cool-water spray at high-risk time points to aid evaporative cooling.

## LIMITATIONS

The demographics of the survey respondents is not a fully representative sample of those participating in the marathon as our sample were predominantly female, of middle age and slower than the average runner. The survey was released seven days after the marathon, increasing the reliance on respondents to accurately recall events from race day and finally, the weather data measured at London, St James’ Park chosen for its central location may not be representative of the weather along the entire course. Future research should aim to capture the responses of a more representative sample of runners, initiate and capture data earlier on, and collect primary data during the marathon using wearable sensors and multiple temperature sensors along the course.

### Conclusion

The 2018 Virgin Money London Marathon was hotter than previous marathons; this subsequently slowed runners finishing time and made runners feel ‘very hot’ and ‘uncomfortable’ for the majority of the race. Our findings have several practical implications, most notably the utilisation of specific analysis on predicted race day temperature and expected finish time as well as the identification of time-points of the race that coincide with increased thermal demand. Integrating a range of strategies highlighted as a result of this research can help to make the race safer and more enjoyable for mass participation runners.

## Data Availability

All data pertaining to the VMLM can be accessed on line via their own website.
Meteorological data can be accessed via the Met Office website

https://www.virginmoneylondonmarathon.com/

